# Neighbourhood-level risk factors of COVID-19 incidence and mortality

**DOI:** 10.1101/2021.01.27.21250618

**Authors:** Trevor van Ingen, Samantha Akingbola, Kevin A. Brown, Nick Daneman, Sarah A. Buchan, Brendan T. Smith

## Abstract

**Background:** Racialized and low income communities face disproportionally high rates of coronavirus 2019 (COVID-19) infection and death. However, data on inequities in COVID-19 across granular categories of socio-demographic characteristics is more sparse.

**Methods:** Neighbourhood-level counts of COVID-19 cases and deaths in Ontario, Canada recorded as of July 28^th^, 2020 were extracted from provincial and local reportable infectious disease surveillance systems. Associations between COVID-19 incidence and mortality and 18 neighbourhood-level measures of immigration, race, housing and socio-economic characteristics were estimated with Poisson generalized linear mixed models. Housing characteristic variables were subsequently added to models to explore if housing may have a confounding influence on the relationships between immigration, race, and socio-economic status and COVID-19 incidence.

**Results:** There were large inequities in COVID-19 incidence and mortality across the socio-demographic variables examined. Neighbourhoods having a higher proportion immigrants, racialized populations, large households and low socio-economic status were associated with COVID-19 risk. Adjusting for housing characteristics, especially unsuitably crowded housing, attenuated COVID-19 risks. However persistent risk remained for neighbourhoods having high proportions of immigrants, racialized populations, and proportion of Black, Latin American, and South Asian residents.

**Conclusions:** Socio-demographic factors account for some of the neighbourhood-level differences in COVID-19 across Ontario. Housing characteristics account for a portion, but not all, of the excess burden of COVID-19 experienced by immigrant, racialized, low income and low education populations.

## Introduction

As the COVID-19 pandemic surges globally, racial and socio-economic inequities in coronavirus 2019 (COVID-19) infections and associated outcomes have emerged as critical public health priorities.[1] Cumulating international evidence has documented disproportionately high rates of COVID-19 infection and mortality experienced by racialized and low income populations. For example, early in the pandemic it was observed that Black adults have four times higher odds of COVID-19 death than White adults in the United Kingdom (UK); South Asian and mixed ethnicity individuals also have significantly higher odds of death.[2] In the United States (US), nearly 70% of early COVID-19 deaths in Chicago, Illinois occurred among Black individuals despite comprising only 30% of the population, with similar patterns observed in other heavily affected areas during the emergence of COVID-19 in the US.[3] Overall, there has been a dearth of US and Canadian COVID-19 surveillance data disaggregated according to important equity stratifiers, such as race and socio-economic status, and when available, has often been incomplete.[4,5] Better understanding the social distribution of COVID-19, particularly within local jurisdictions where COVID-19 trends differ, is critical towards informing the design of equitable policy and intervention strategies to reduce the burden of COVID-19.

Area-level measures of socio-economic status are commonly used to better understand inequities in COVID-19 incidence and mortality when individual-level measures are not available. Ecological studies using area-level measures have revealed widespread evidence of social inequities in COVID-19 incidence and mortality associated with race, poverty, and residential crowding.[6–11] While these trends have been consistently identified, the specific populations at risk and strength of association vary between jurisdictions. With limited data available in Canada, evidence is urgently needed to understand the extent to which COVID-19 outcomes and risk factors vary according to race and socioeconomic status in a Canadian context.

In Ontario – Canada’s largest province by population – inequities in the burden of COVID-19 among both ethno-racially diverse and low socioeconomic neighbourhoods have been identified.[12–14] While these findings confirm that socio-demographic factors influence COVID-19 risk in Ontario, they lack the ability to disaggregate findings by granular race and socio-economic categories. Further, these analyses do not consider how housing characteristics, especially household size and overcrowding, may contribute to the higher impact of COVID-19 in racialized and low socioeconomic communities. Therefore, the objective of this study was to estimate the associations between neighbourhood socio-demographic characteristics and neighbourhood-level incidence and mortality of COVID-19 in Ontario. Of primary interest was to understand inequities across more granular categories of immigration, race, housing, and socio-economic characteristics. A secondary objective was to evaluate if the association between neighbourhood proportion of immigration, race, and socio-economic status and COVID-19 incidence was independent of household characteristics.

## Methods

### Study Design and Population

We conducted a cross-sectional study using data extracted from provincial and local reportable infectious disease surveillance systems (collectively known as CCM plus) which include all known COVID-19 infections and deaths from Ontario, Canada reported between January 23, 2020 and July 28, 2020.

The study population includes all Ontario community residents reported to CCM plus who met the provincial case definition for COVID-19 (i.e., positive nucleic acid amplification test). Due to relatively poor representativeness of people living in institutions, congregate living settings, and Indigenous communities living on reserves in the Canadian Census (from which exposure and denominator data are derived), these populations were excluded from this study. Residents of institutional and congregate living settings were identified and removed from our sample if they were flagged as such in CCM plus, or their residential address was matched to a comprehensive list of addresses of known institutions and congregate settings using a natural language processing algorithm. These settings and addresses include long-term care facilities, retirement homes, shelters, correction or detention centres, hotels and motels, group homes, hospitals, and on-site accommodations for farm workers.

### Covariables

Neighbourhood socio-demographic characteristics were derived from the 2016 Census of Population using Statistics Canada Aggregate Dissemination Areas (ADA) as the measure for neighbourhood. Neighbourhoods with small populations (<1,000 people) were excluded due to stability concerns. On average, neighbourhood populations ranged between 5,000 to 15,000 people. Case records from CCM plus were assigned to a neighbourhood based on postal code of residence using Statistics Canada Postal Code Conversion File (PCCF) plus version 7C. Eighteen neighbourhood-level socio-demographic measures were included in this study, including: 1) eight measures of the proportion of immigration and race (immigrants, recent immigrants (within five years), visible minority (non-white and non-Indigenous population), Black, East/Southeast Asian, Latin American, Middle Eastern, and South Asian); 2) six measures of housing characteristics (average household size, proportion multigenerational families (households with at least one person living with a child and a grandparent), proportion unsuitably crowded housing (according to the National Occupancy Standard on housing suitability based on number of bedrooms and composition of the household), proportion of dwellings in apartments in flat/duplex, proportion of dwellings in low-rise apartments (≤4 storey), and proportion of dwellings in high-rise apartments (≥5 storeys)); and 3) four socio-economic status measures (labour force participation (age ≥15 years), proportion without a high school diploma (age ≥25 years), proportion low income (measured using after-tax low income cut-offs (LICO)), and proportion unaffordable housing (spending >30% of income on housing)). Urban/rural geographic stratification was determined by grouping neighbourhoods into four categories based on community size, population density, and level of integration with a census metropolitan area or census agglomeration.[15]

### Outcomes

Cumulative (until July 28, 2020) incidence and mortality rates were calculated using the 2016 census population denominators, as more recent population projections were not available at the neighbourhood-level. Postal code of residence, age group (<15, 15-64, and ≥65 years), sex (male, female), case status, and outcome status were extracted from CCM plus.

This study received ethics approval from Public Health Ontario’s Research Ethics Board.

### Statistical Analysis

Associations between neighbourhood-level characteristics and neighbourhood-level counts of COVID-19 infections and deaths were estimated by running a series of Poisson generalized linear mixed models with random effects for neighbourhood, offset for neighbourhood population, using the ‘glmer’ package in R. Bivariable models were used to estimate associations between neighbourhood measures of immigration, race, housing, and socio-economic status and COVID-19 incidence and mortality. Subsequent multivariable models were adjusted for individual-level age, sex, and neighbourhood urban/rural geography. Model estimates were standardized to show relative risks and 95% confidence intervals of COVID-19 incidence and mortality between the 10^th^ (p10) and 90^th^ (p90) percentile of each neighbourhood socio-demographic characteristic.

To explore how housing characteristics may confound relationships between COVID-19 incidence and neighbourhood-level immigration, race and socio-economic characteristics, an additional series of multivariable models adjusting for housing characteristic variables (average household size, proportion multigenerational families, and proportion unsuitably crowded housing) were run.

## Results

Between January 23^rd^ and July 28^th^, 38,984 individuals with confirmed COVID-19 were recorded in CCM plus in Ontario. Of those, 37,343 individuals (96%) had a valid postal code record that was successfully assigned to a neighbourhood. A further 8,822 (24%) residents of congregate settings, and 59 (0.5%) Indigenous communities living on reserves or small neighbourhoods with populations <1,000 were excluded. In total, our study population included 28,808 people with COVID-19 infection and 683 COVID-19 deaths. Socio-demographic data were derived for 1,526 Ontario neighbourhoods.

### COVID-19 incidence and mortality across neighbourhoods

The distribution of COVID-19 incidence and mortality varied across neighbourhoods in Ontario (Figure 1). The top 10% highest incidence neighbourhoods accounted for 36% of the cases. The highest rate of COVID-19 incidence was 1,771 per 100,000. In nearly 70% of neighbourhoods, there were zero COVID-19 deaths and the top 10% highest mortality neighbourhoods accounted for 59% of all COVID-19 deaths. The highest neighbourhood COVID-19 mortality rate was 96 per 100,000.

**Figure 1.**
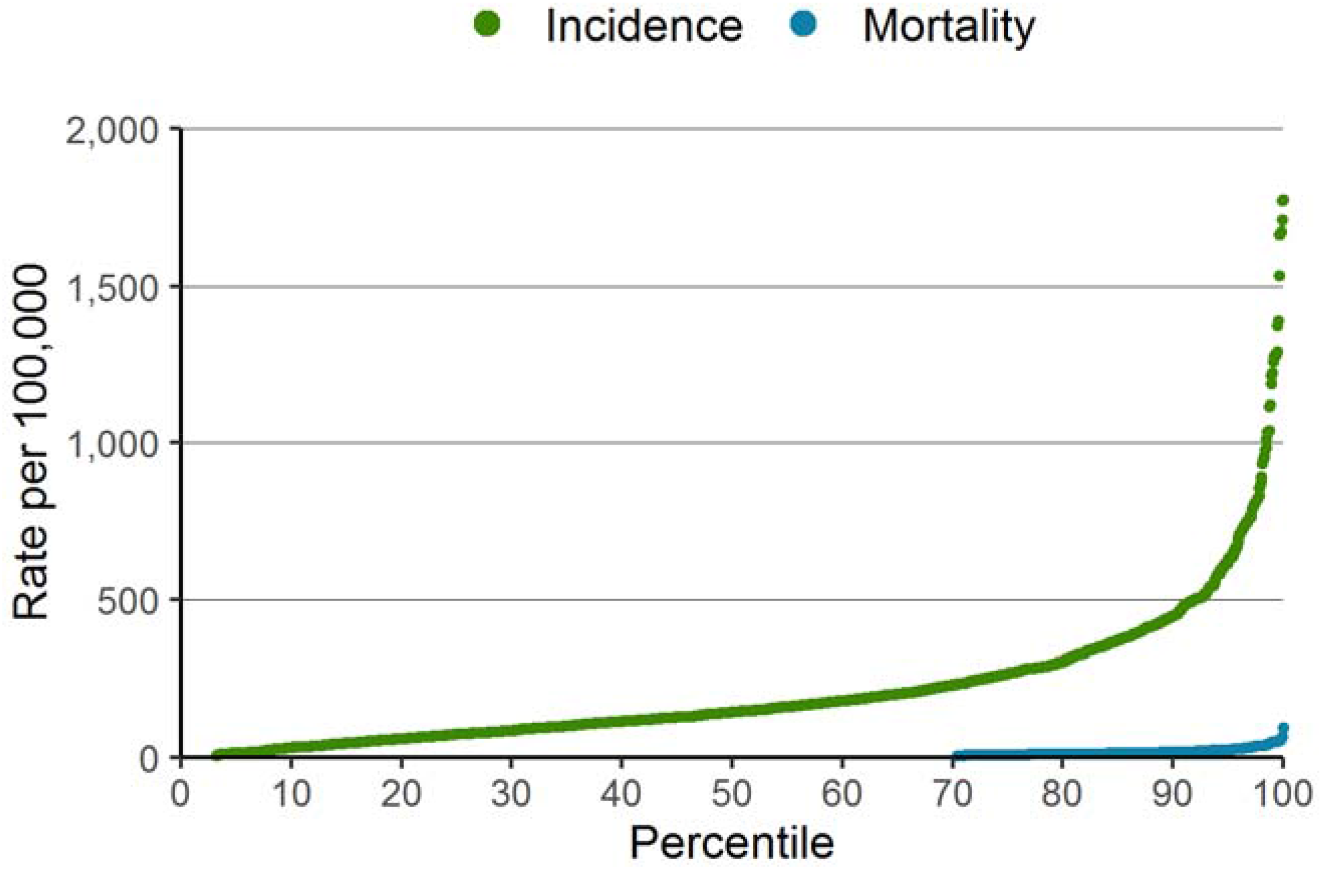
Neighbourhood incidence and mortality rate per 100,000, ranked by neighbourhood percentile

**Figure 2a.**
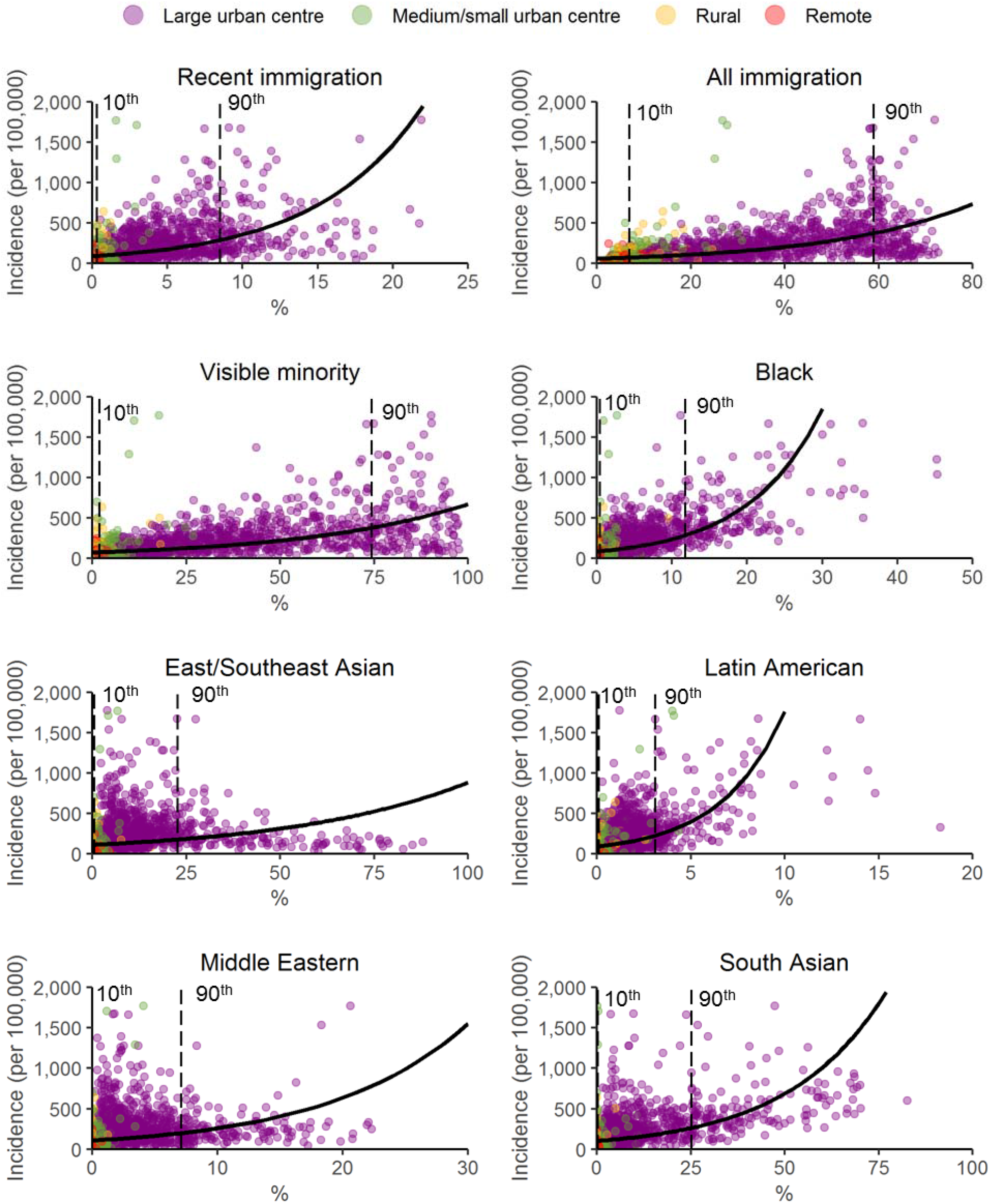
Neighbourhood-level incidence of COVID-19 by proportion of immigrant and race, with regression estimates and 10^th^ and 90^th^ percentiles.

**Figure 2b.**
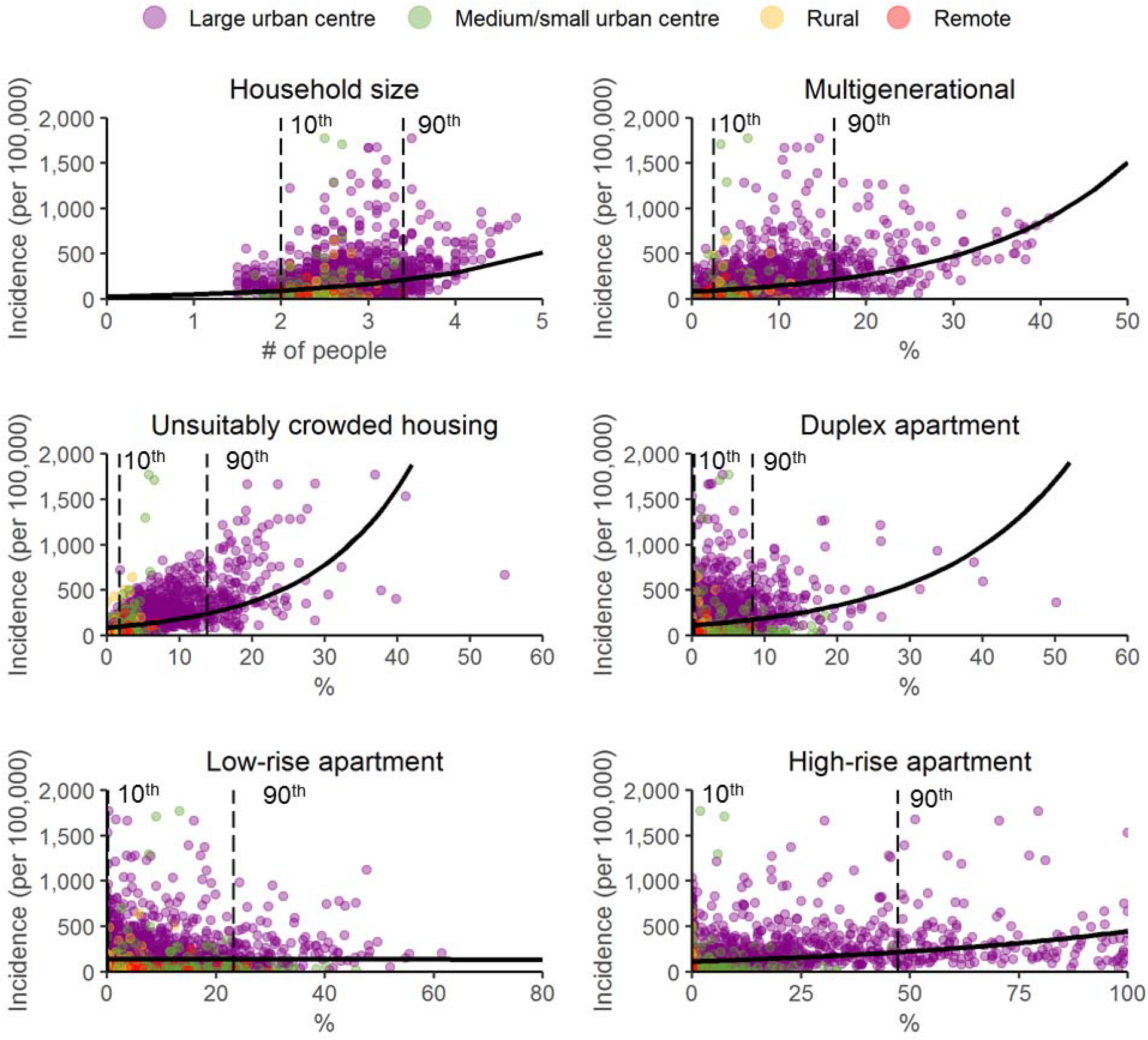
Neighbourhood-level incidence of COVID-19 by proportion of housing characteristics, with regression estimates and 10^th^ and 90^th^ percentiles.

**Figure 2c.**
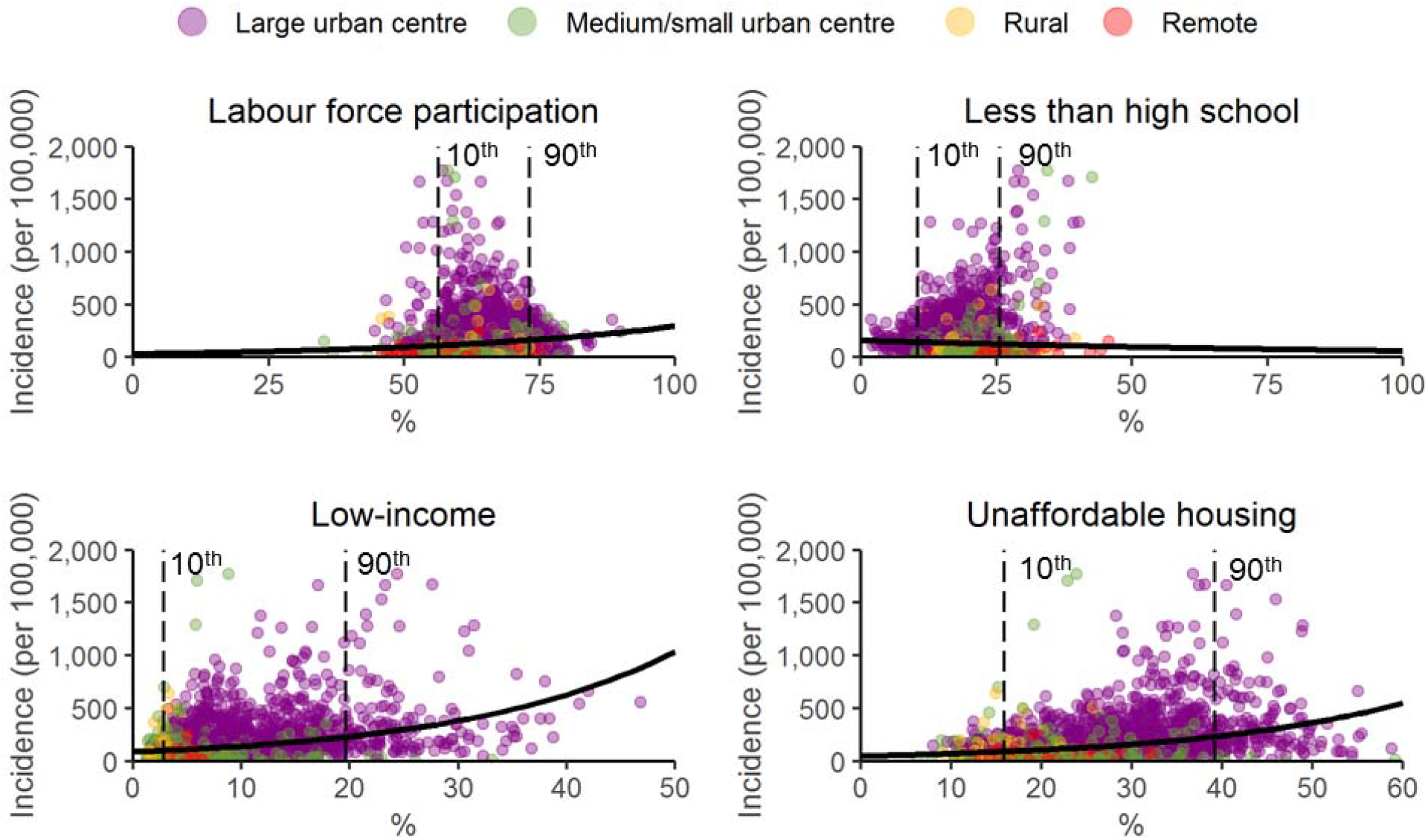
Neighbourhood-level incidence of COVID-19 by proportion of socio-economic status characteristics, with regression estimates and 10 ^th^ and 90 ^th^ percentiles.

The 18 neighbourhood-level measures we explored are described in terms of composition of the neighbourhoods (neighbourhood median and interdecile range), and incidence and mortality for neighbourhoods in the lowest and highest decile of that characteristic (Table 1). For most neighbourhood characteristics, the rates of COVID-19 incidence and mortality were higher among the highest compared to lowest decile, with the exception for neighbourhood low-rise apartments, and neighbourhood labour force participation. COVID-19 incidence was highest among the neighbourhoods in the highest decile of proportion Black residents, and mortality was highest among the neighbourhoods in the highest decile of unsuitably crowded housing.

**Table 1.** Median neighbourhood socio-demographic characteristics, interdecile range, and incidence and mortality of COVID-19 in the lowest and highest deciles of a given neighbourhood characteristic

| Characteristic | Median<br>(interdecile range) | COVID-19 Incidence |  | COVID-19 Mortality |  |
| --- | --- | --- | --- | --- | --- |
|  |  | Lowest decile | Highest decile | Lowest decile | Highest decile |
| Cases (Incidence per 100,000) |  |  |  |  |  |
| Immigration and race |  |  |  |  |  |
| All immigrants (%) | 24.9 (6.9 – 59.0) | 683 (54.8) | 5,312 (392.1) | 14 (1.1) | 143 (10.6) |
| Recent immigrant (%) | 2.4 (0.3 – 8.5) | 818 (67.8) | 6,039 (434.0) | 24 (2.0) | 161 (11.6) |
| Visible Minority Status (%) | 20.2 (1.9 – 74.4) | 735 (59.9) | 6,754 (473.1) | 16 (1.3) | 141 (9.9) |
| Black (%) | 2.7 (0.4 – 11.8) | 849 (68.5) | 8,012 (583.7) | 18 (1.5) | 152 (11.1) |
| East/Southeast Asian (%) | 5.6 (0.6 – 22.7) | 912 (74.7) | 3,398 (243.0) | 20 (1.6) | 105 (7.5) |
| Latin American (%) | 1.1 (0.1 – 3.1) | 797 (65.5) | 7,111 (510.5) | 20 (1.6) | 157 (11.3) |
| Middle Eastern (%) | 1.4 (0.0 – 7.1) | 1,100 (66.4) | 3,705 (270.2) | 31 (1.9) | 93 (6.8) |
| South Asian (%) | 3.4 (0.3 – 25.2) | 975 (82.1) | 7,146 (506.9) | 28 (2.4) | 145 (10.3) |
| Housing |  |  |  |  |  |
| Average household size (N) | 2.6 (2.1 – 3.5) | 2,902 (157.1) | 4,514 (413.6) | 89 (4.8) | 76 (7.0) |
| Multigenerational families (%) | 6.1 (2.5 – 16.3) | 1,700 (126.1) | 6,169 (460.6) | 43 (3.2) | 107 (8.0) |
| Unsuitably crowded housing (%) | 3.9 (1.7 – 13.8) | 935 (75.9) | 8,019 (566.8) | 24 (1.9) | 194 (13.7) |
| Apartment in duplex or flat (%) | 2.0 (0.3 – 8.3) | 2,539 (188.2) | 4,151 (333.3) | 65 (4.8) | 91 (7.3) |
| Low-rise apartment (%) | 6.0 (0.2 – 23.2) | 3,383 (252.9) | 2,965 (219.4) | 77 (5.8) | 72 (5.3) |
| High-rise apartment (%) | 3.8 (0.0 – 47.3) | 7,815 (170.9) | 5,257 (357.4) | 157 (3.4) | 166 (11.3) |
| Socio-economic status |  |  |  |  |  |
| Labour force participation (%) | 64.9 (56.3 – 73.1) | 2,471 (190.2) | 2,547 (171.8) | 69 (5.3) | 42 (2.8) |
| Less than high school (%) | 17.0 (10.4 – 25.6) | 2,295 (155.8) | 4,721 (377.7) | 81 (5.5) | 98 (7.8) |
| Low income (%) | 7.7 (2.8 – 19.6) | 1,301 (97.8) | 5,333 (388.1) | 29 (2.2) | 144 (10.5) |
| Unaffordable housing (%) | 25.9 (15.9 – 39.2) | 1,436 (113) | 5,385 (363.3) | 32 (2.5) | 157 (10.6) |

### Modeling estimates

The rate of COVID-19 incidence per 100,000 are plotted as a function of neighbourhood-level immigration and race (Figure 2a), housing (Figure 2b), and socio-economic status (Figure 2c), with bivariable regression estimates and lines marking the 10^th^ and 90^th^ percentile of each predictor’s distribution. Across 16 of the 18 predictors, as the proportion of the neighbourhood-level characteristic increases, so did the incidence of COVID-19. There was no association with proportion of low-rise apartments, and as neighbourhood proportion with less than high school education increases, COVID-19 decreases.

COVID-19 incidence and mortality risk were estimated using both bivariable and multivariable models (Table 3). Overall, our models estimate that neighbourhoods in the 90^th^ percentile (p90) of socio-demographic characteristics were associated with higher rates of COVID-19 incidence and mortality compared to neighbourhoods with in the 10^th^ percentile (p10) of socio-demographic characteristics for most unadjusted models. Adjusting for age, sex, and urban/rural geographies reduced the strength of most associations.

**Table 3.** Relative risks of COVID-19 incidence between 10 ^th^ and 90 ^th^ percentile of neighbourhood-level proportion of socio-demographic characteristics.

| Characteristic | Incidence<br>relative risk –<br>p10 vs p90<br>(95% CI) | Adjusted*<br>incidence<br>relative risk –<br>p10 vs p90<br>(95% CI) | Mortality<br>relative risk –<br>p10 vs p90<br>(95% CI) | Adjusted*<br>mortality<br>relative risk –<br>p10 vs p90<br>(95% CI) |
| --- | --- | --- | --- | --- |
| **Immigration and race** |  |  |  |  |
| Recent immigrant | 3.2 (2.9 – 3.5) | 2.1 (1.9 – 2.4) | 2.8 (2.3 – 3.4) | 2.5 (2.0 – 3.1) |
| All immigrants | 5.4 (4.9 – 5.9) | 4.0 (3.5 – 4.5) | 5.2 (4.0 – 6.7) | 5.2 (3.8 – 7.1) |
| Visible Minority Status | 5.0 (4.5 – 5.5) | 3.3 (2.9 – 3.7) | 3.7 (2.9 – 4.8) | 3.5 (2.6 – 4.6) |
| Black | 3.2 (3.0 – 3.4) | 2.4 (2.2 – 2.6) | 2.2 (1.9 – 2.5) | 2.0 (1.7 – 2.3) |
| East/Southeast Asian | 1.6 (1.5 – 1.7) | 1.0 (1.0 – 1.1) | 1.5 (1.3 – 1.8) | 1.2 (1.0 – 1.4) |
| Latin American | 2.4 (2.3 – 2.6) | 1.8 (1.7 – 1.9) | 1.9 (1.6 – 2.1) | 1.6 (1.4 – 1.8) |
| Middle Eastern | 1.9 (1.7 – 2.1) | 1.2 (1.1 – 1.3) | 1.6 (1.3 – 1.9) | 1.3 (1.0 – 1.5) |
| South Asian | 2.6 (2.4 – 2.8) | 1.9 (1.8 – 2.1) | 1.9 (1.6 – 2.2) | 1.8 (1.5 – 2.1) |
| **Housing** |  |  |  |  |
| Average household size | 2.2 (2.0 – 2.5) | 1.9 (1.7 – 2.1) | 1.5 (1.1 – 1.9) | 1.6 (1.2 – 2.1) |
| Multigenerational families | 2.2 (2.0 – 2.4) | 1.8 (1.7 – 2.0) | 1.8 (1.5 – 2.2) | 1.7 (1.4 – 2.1) |
| Unsuitably crowded housing | 2.4 (2.3 – 2.6) | 2.1 (2.0 – 2.3) | 2.5 (2.2 – 2.9) | 2.5 (2.1 – 2.9) |
| Apartment in duplex or flat | 1.5 (1.4 – 1.7) | 1.3 (1.2 – 1.4) | 1.4 (1.2 – 1.6) | 1.2 (1.0 – 1.4) |
| Low-rise apartment | 1.0 (0.9 – 1.1) | 0.9 (0.8 – 1.0) | 1.1 (0.9 – 1.4) | 1.0 (0.8 – 1.2) |
| High-rise apartment | 1.9 (1.8 – 2.1) | 1.2 (1.1 – 1.4) | 2.3 (1.9 – 2.7) | 1.7 (1.4 – 2.1) |
| **Socio-economic status** |  |  |  |  |
| Labour force participation | 1.5 (1.3 – 1.6) | 1.0 (0.9 – 1.1) | 0.7 (0.5 – 0.9) | 0.8 (0.6 – 1.0) |
| Less than high school education | 0.8 (0.8 – 0.9) | 1.6 (1.5 – 1.8) | 1.3 (1.1 – 1.7) | 2.1 (1.6 – 2.7) |
| Low income | 2.1 (1.9 – 2.3) | 1.4 (1.2 – 1.5) | 2.3 (1.9 – 2.8) | 1.8 (1.5 – 2.2) |
| Unaffordable housing | 2.6 (2.3 – 2.9) | 1.6 (1.4 – 1.8) | 3.1 (2.4 – 3.9) | 2.4 (1.8 – 3.1) |

\* Adjusted for age-group, sex, and urban/rural stratifier

Neighbourhoods with high labour force participation had lower rates of COVID-19 mortality compared to neighbourhoods with low labour force participation. The proportion of low-rise apartments was not associated with COVID-19 incidence or mortality (Table 3). Fully adjusted models also reversed the protective association between neighbourhoods with the highest compared to lowest proportion of less than high school education and COVID-19 incidence, controlling for confounding related to urban/rural geographies.

The proportion of immigrants in a neighbourhood showed the strongest association with COVID-19, with an incidence relative risk of 4.0 (95% CI: 3.5 – 4.5) and mortality relative risk of 5.2 (95% CI: 3.8 – 7.1) in fully adjusted models. The proportion of visible minority residents showed stronger associations with incidence and mortality compared to the proportion of residents from individual race groups. Neighbourhood proportions of Black residents, followed by proportion South Asian residents, and proportion Latin American residents show the strongest associations of individual race groups.

### Adjusting for housing characteristics

Exploratory models that added housing characteristic terms to models of immigration, race and socio-economic characteristics and COVID-19 incidence were run with adjustments for age-group, sex, and urban/rural geography (Table 4). In most cases, adding housing characteristic predictors reduced the association between immigration, race, and socio-economic status and COVID-19 incidence.

**Table 4.**
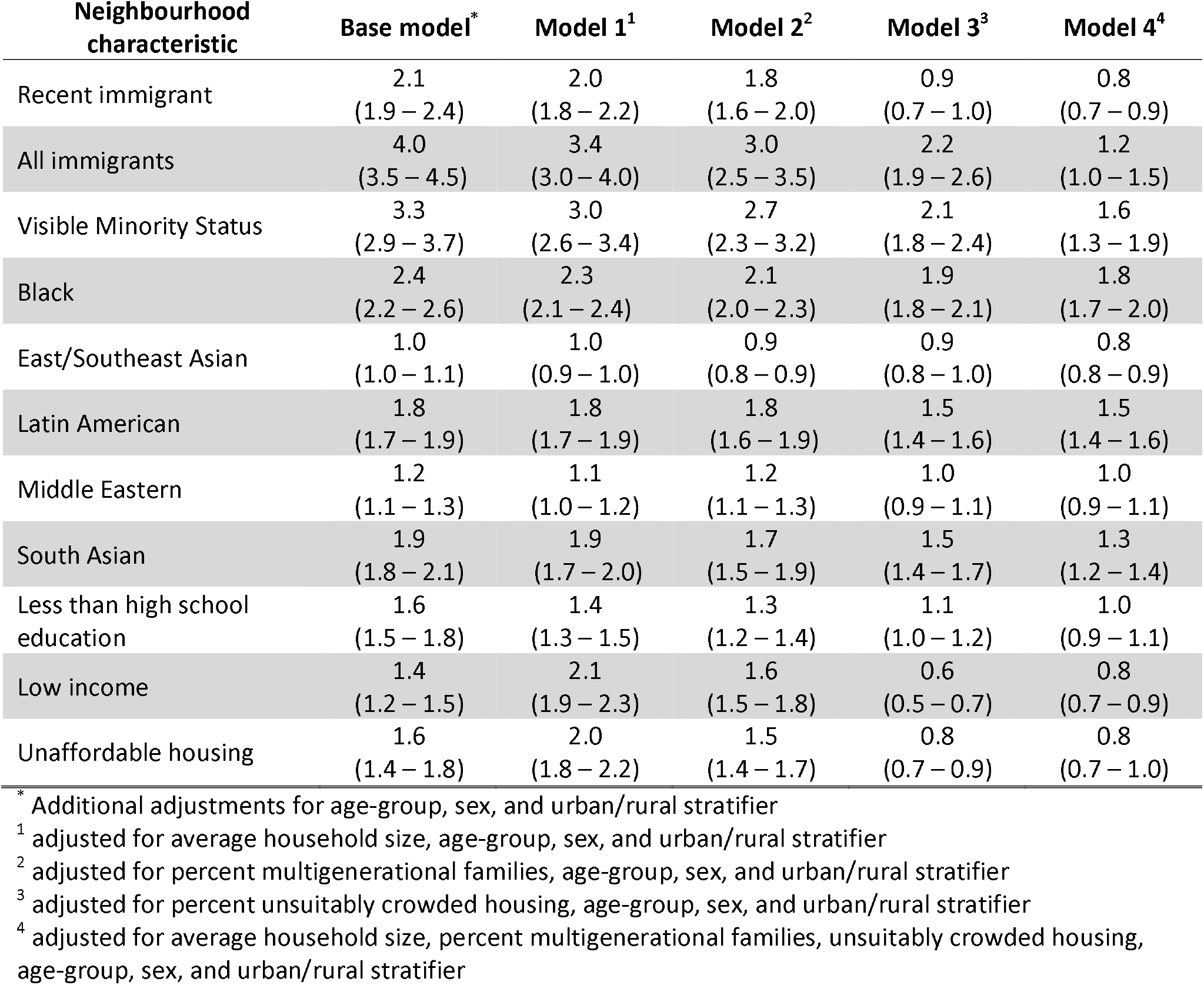
Relative risks of COVID-19 incidence between 10 ^th^ and 90 ^th^ percentile of neighbourhood-level proportion of race and immigration characteristics, adjusted by household characteristics.

| Neighbourhood characteristic | Base model<sup>*</sup> | Model 1<sup>1</sup> | Model 2<sup>2</sup> | Model 3<sup>3</sup> | Model 4<sup>4</sup> |
| --- | --- | --- | --- | --- | --- |
| Recent immigrant | 2.1<br>(1.9 – 2.4) | 2.0<br>(1.8 – 2.2) | 1.8<br>(1.6 – 2.0) | 0.9<br>(0.7 – 1.0) | 0.8<br>(0.7 – 0.9) |
| All immigrants | 4.0<br>(3.5 – 4.5) | 3.4<br>(3.0 – 4.0) | 3.0<br>(2.5 – 3.5) | 2.2<br>(1.9 – 2.6) | 1.2<br>(1.0 – 1.5) |
| Visible Minority Status | 3.3<br>(2.9 – 3.7) | 3.0<br>(2.6 – 3.4) | 2.7<br>(2.3 – 3.2) | 2.1<br>(1.8 – 2.4) | 1.6<br>(1.3 – 1.9) |
| Black | 2.4<br>(2.2 – 2.6) | 2.3<br>(2.1 – 2.4) | 2.1<br>(2.0 – 2.3) | 1.9<br>(1.8 – 2.1) | 1.8<br>(1.7 – 2.0) |
| East/Southeast Asian | 1.0<br>(1.0 – 1.1) | 1.0<br>(0.9 – 1.0) | 0.9<br>(0.8 – 0.9) | 0.9<br>(0.8 – 1.0) | 0.8<br>(0.8 – 0.9) |
| Latin American | 1.8<br>(1.7 – 1.9) | 1.8<br>(1.7 – 1.9) | 1.8<br>(1.6 – 1.9) | 1.5<br>(1.4 – 1.6) | 1.5<br>(1.4 – 1.6) |
| Middle Eastern | 1.2<br>(1.1 – 1.3) | 1.1<br>(1.0 – 1.2) | 1.2<br>(1.1 – 1.3) | 1.0<br>(0.9 – 1.1) | 1.0<br>(0.9 – 1.1) |
| South Asian | 1.9<br>(1.8 – 2.1) | 1.9<br>(1.7 – 2.0) | 1.7<br>(1.5 – 1.9) | 1.5<br>(1.4 – 1.7) | 1.3<br>(1.2 – 1.4) |
| Less than high school education | 1.6<br>(1.5 – 1.8) | 1.4<br>(1.3 – 1.5) | 1.3<br>(1.2 – 1.4) | 1.1<br>(1.0 – 1.2) | 1.0<br>(0.9 – 1.1) |
| Low income | 1.4<br>(1.2 – 1.5) | 2.1<br>(1.9 – 2.3) | 1.6<br>(1.5 – 1.8) | 0.6<br>(0.5 – 0.7) | 0.8<br>(0.7 – 0.9) |
| Unaffordable housing | 1.6<br>(1.4 – 1.8) | 2.0<br>(1.8 – 2.2) | 1.5<br>(1.4 – 1.7) | 0.8<br>(0.7 – 0.9) | 0.8<br>(0.7 – 1.0) |

<sup>\*</sup> Additional adjustments for age-group, sex, and urban/rural stratifier
<sup>1</sup> adjusted for average household size, age-group, sex, and urban/rural stratifier
<sup>2</sup> adjusted for percent multigenerational families, age-group, sex, and urban/rural stratifier
<sup>3</sup> adjusted for percent unsuitably crowded housing, age-group, sex, and urban/rural stratifier
<sup>4</sup> adjusted for average household size, percent multigenerational families, unsuitably crowded housing, age-group, sex, and urban/rural stratifier

Of the housing characteristics, adjusting for unsuitably crowded housing had the greatest impact on these associations. The association of neighbourhood proportion immigrants, visible minority status, proportion of Black, Latin American, and South Asian residents and COVID-19 incidence were attenuated, but persistent risk remained after adjusting for all household characteristics. However, these adjustments produced null associations for neighbourhood proportion recent immigrants, Middle Eastern, East/Southeast Asian, low income, less than high school education, and unaffordable housing.

## Discussion

Linking COVID-19 surveillance data to neighbourhood-level characteristics from Ontario, Canada, this study found higher COVID-19 incidence and mortality rates in neighbourhoods with a higher proportion of immigrants, racialized populations, large households and low socio-economic status. These findings highlight how structural barriers are acting as key determinants of COVID-19 inequities. Additionally, housing characteristics, in particular unsuitably crowded housing, are important factors increasing COVID-19 risk among immigrant, racialized and low socio-economic communities.

People living in the most marginalized neighbourhoods are experiencing elevated rates of COVID-19 outcomes, both in Canada[12,13] and internationally.[6,7,11,16–18] For example, US counties with the highest compared to the lowest proportion of populations of colour and poverty had 4.9 and 1.7 times higher rates of COVID-19 death.[7] Similarly in Canada, age-standardized COVID-19 mortality rates were two times higher in neighbourhoods with the highest (>25%) compared to lowest (<1%) proportion of visible minorities, although this varied by province with COVID-19 mortality rates 3.4 higher in Ontario.[19] Our study adds to this body of evidence by examining associations between specific socio-demographic characteristics and COVID-19 burden. We found neighbourhoods with a high proportion of immigrants had four times higher risk of COVID-19 infection and 5.2 times higher risk of death, followed by neighbourhoods with a high proportion of visible minority residents which had 3.3 times higher risk of COVID-19 infection and 3.5 times higher risk of death. Further, the increased risk among neighbourhoods with high proportions of Black, South Asian, and Latin American populations found in our study are consistent with a a recent systematic-review and meta-analysis of 50 studies on individual-level ethnicity and COVID-19 from the UK and US.[20]

Existing and persistent structural inequities put immigrant, racialized, and low-income communities at higher risk of COVID-19 exposure and infection. Housing conditions, especially household size and crowding, is an important predictor of COVID-19 transmission.[7,21,22] In Canada, 21.1% of racialized individuals live in unsuitably crowded households, nearly four times the rate of non-racialized individuals.[23] Additionally, immigrants are twice as likely to live in multigenerational households compared to non-immigrants.[24] In our study, adjusting for measures of housing characteristics attenuated the associations but social inequities in COVID-19 incidence remained, suggesting they play an important role but do not fully explain the disproportionate impact of COVID-19 on these communities.

Low socio-economic status further accounts for excess COVID-19 burden experienced by immigrant and racialized populations, impacting the ability to avoid COVID-19 infection at work. Evidence from the literature describes significant risks of COVID-19 associated with occupations, especially those in precarious employment, where it is difficult to distance from others, there is increased risk of exposure to infections while at work, and employers are less likely to provide paid sick leave.[25–28] In Canada, immigrant and racialized individuals are not only more likely to live in poverty,[29] but also overrepresented in risky essential occupations. For example, immigrants and racialized populations in Ontario are disproportionately employed in long-term care facilities as nurse aids, orderlies, and patient service associates.[30] Further, COVID-19 testing data from Ontario confirmed that a disproportionate number of immigrants diagnosed with COVID-19 were employed as health care workers.[31] Further exploring the role that occupation and workplace settings has on contributing to inequities in COVID-19 transmission and negative outcomes in Ontario represents an important area of future study.

Other explanations for the stark inequities in COVID-19 outcomes found in this study may be rooted in systematic barriers faced by racialized and newcomer populations.[32] In Canada, racism and other structural determinants are an underlying cause of the overrepresentation of Black individuals having lower socio-economic status and inadequate access to a regular doctor.[33,34] Racialized populations also experience pre-existing health inequities, such as higher rates of comorbidities, which could also contribute to greater vulnerability to severe COVID-19 outcomes.[35,36] However, data from the first wave in Ontario showed that although incidence and mortality were higher among diverse neighbourhoods, the case-fatality ratio was lower, suggesting that greater number of infections, and not co-morbidities, was a driving cause for the increased rate of death.[12] Systemic racism has been observed in other Western jurisdictions in ways that can contribute to increased COVID-19 risk. In the US, racialized individuals are more likely to be incarcerated, which in turn increases the likelihood of infection,[8] and racial discrimination in mortgage lending practices in urban areas may be a driving factor in residential crowding among Black Americans.[37] In the UK, non-White physicians comprise the vast majority of COVID-19 deaths among doctors and, compared to White physicians, are more likely to report being under pressure to attend to patients without receiving the necessary physical protection.[38]

This study is subject to some limitations. First, the number of COVID-19 cases included in this study is an undercount of the true number of infections, and may be biased by changes in testing criteria during the study period and differences in testing patterns between various socio-demographic populations. However, recent Ontario research describing both the odds of having been tested for COVID-19 and the odds of having received a positive COVID-19 diagnosis suggest that selection bias is not driving socio-demographic inequities in COVID-19 incidence in the province.[39] Additionally, the use of neighbourhood-level measures of socio-demographic characteristics may dilute the real effect of these characteristics on COVID-19 risks, as individual cases may not reflect the characteristics of the neighbourhoods they live in. Previous Canadian studies comparing individual and area-level measures have shown that even with relatively poor agreement between measures, area-level measures may be describing important community-level effects that contribute to health inequities.[40] The collection of individual-level socio-demographic data in Ontario during subsequent phases of the pandemic will allow for future validation of our findings.

This study has several strengths. The ability to use postal code to link people with COVID-19 to census data at the neighbourhood-level provides greater accuracy than is possible with most publically available data on COVID-19. Additionally, we included a large number of socio-demographic characteristics, notably the ability to examine specific immigration, race, housing and socio-economic categories. Finally, this is the first study to demonstrate the impact that neighbourhood-level household characteristics have on the associations between immigration, race, income and education and COVID-19 incidence in Ontario. By including housing factors in our models, we have confirmed the importance of housing as a focus for public health interventions.

## Conclusion

Neighbourhood socio-demographic factors appear to explain much of the neighbourhood-level variability in COVID-19 across Ontario. Housing characteristics, especially unsuitably crowded housing, seem to account for a portion of the burden of COVID-19 experienced by immigrant, racialized, low income and low education populations. These results suggest that prevention of household transmission, in addition to culturally safe approaches to engaging with racialized communities and communities living in poverty, are important public health strategies for reducing COVID-19 inequities. Future research on COVID-19 inequities should focus how the relationship between the socio-demographic factors examined in this study and COVID-19 are confounded by occupation and workplace characteristics.

## Data Availability

Data sharing requests should be directed to Public Health Ontario.

## References

1 Pareek M, Bangash MN, Pareek N, et al. Ethnicity and COVID-19: an urgent public health research priority. The Lancet 2020;395:1421–2. doi:10.1016/S0140-6736(20)30922-3

2 Office for National Statistics. Coronavirus (COVID-19) related deaths by ethnic group, England and Wales. 2020.https://www.ons.gov.uk/peoplepopulationandcommunity/birthsdeathsandmarriages/deaths/articles/coronavirusrelateddeathsbyethnicgroupenglandandwales/2march2020to10april2020 (accessed 12 Nov 2020).

3 Yancy CW. COVID-19 and African Americans. JAMA 2020;323:1891. doi:10.1001/jama.2020.6548

4 Blair A, Warsame K, Naik H, et al. Identifying gaps in COVID-19 health equity data reporting in Canada using a scorecard approach. MedRxiv 20200147 [Preprint]. 25 September 2020. [cited 2020 Oct 21] doi:10.1101/2020.09.23.20200147

5 Krieger N, Gonsalves G, Bassett MT, et al. The Fierce Urgency Of Now: Closing Glaring Gaps In US Surveillance Data On COVID-19. Health Affairs Blog. 2020. doi:10.1377/hblog20200414.238084

6 Adhikari S, Pantaleo NP, Feldman JM, et al. Assessment of Community-Level Disparities in Coronavirus Disease 2019 (COVID-19) Infections and Deaths in Large US Metropolitan Areas. JAMA Netw Open 2020;3:e2016938. doi:10.1001/jamanetworkopen.2020.16938

7 Chen JT, Krieger N. Revealing the Unequal Burden of COVID-19 by Income, Race/Ethnicity, and Household Crowding: US County Versus Zip Code Analyses. Journal of Public Health Management and Practice 2020;27:S43–56. doi:10.1097/PHH.0000000000001263

8 Chin T, Kahn R, Li R, et al. US-county level variation in intersecting individual, household and community characteristics relevant to COVID-19 and planning an equitable response: a cross-sectional analysis. BMJ Open 2020;10:e039886. doi:10.1136/bmjopen-2020-039886

9 Hawkins D. Social Determinants of COVID-19 in Massachusetts, United States: An Ecological Study. J Prev Med Public Health 2020;53:220–7. doi:10.3961/jpmph.20.256

10 Plümper T, Neumayer E. The pandemic predominantly hits poor neighbourhoods? SARS-CoV-2 infections and COVID-19 fatalities in German districts. European Journal of Public Health 2020;30:1176–80. doi:10.1093/eurpub/ckaa168

11 Wadhera RK, Wadhera P, Gaba P, et al. Variation in COVID-19 Hospitalizations and Deaths Across New York City Boroughs. JAMA 2020;323:2192. doi:10.1001/jama.2020.7197

12 Ontario Agency for Health Protection and Promotion (Public Health Ontario). COVID-19 in Ontario - A focus on diversity: January 15, 2020 to May 14, 2020. 2020.https://www.publichealthontario.ca/-/media/documents/ncov/epi/2020/06/covid-19-epi-diversity.pdf?la=en (accessed 3 Nov 2020).

13 Ontario Agency for Health Protection and Promotion (Public Health Ontario). COVID-19 in Ontario - A focus on material deprivation: January 15, 2020 to June 3, 2020. 2020.https://www.publichealthontario.ca/-/media/documents/ncov/epi/2020/06/covid-19-epi-material-deprivation.pdf?la=en (accessed 3 Nov 2020).

14 Chung H, Fung K, Ferreira-Legere L, et al. COVID-19 Laboratory Testing in Ontario: Patterns of Testing and Characteristics of Individuals Tested, as of April 30, 2020. 2020.https://www.ices.on.ca/~/media/Files/Atlases-Reports/2020/COVID-19-Laboratory-Testing-in-Ontario/Full-report.ashx (accessed 13 Nov 2020).

15 Health Quality Ontario. Geographic Location Methods Review: Summary Report. 2019.https://www.hqontario.ca/Portals/0/documents/pr/hqo-geographic-location-methods-review-report.pdf (accessed 15 Nov 2020).

16 Gross CP, Essien UR, Pasha S, et al. Racial and Ethnic Disparities in Population-Level Covid-19 Mortality. J GEN INTERN MED 2020;35:3097–9. doi:10.1007/s11606-020-06081-w

17 Guijarro C, Pérez-Fernández E, Stat M, et al. Differential risk for COVID-19 in the first wave of the disease among Spaniards and migrants from different areas of the world living in Spain. Revista Clínica Española 2020;:14. doi:10.1016/j.rce.2020.10.006

18 Ingraham NE, Purcell LN, Karam BS, et al. Racial/Ethnic Disparities in Hospital Admissions from COVID-19 and Determining the Impact of Neighborhood Deprivation and Primary Language. MedRxiv 20185983 [Preprint]. September 22, 2020 [cited 2020 Oct 21] doi:10.1101/2020.09.02.20185983

19 Subedi R, Greenberg L, Turcotte M. COVID-19 mortality rates in Canada’s ethno-cultural neighbourhoods. https://www150.statcan.gc.ca/n1/pub/45-28-0001/2020001/article/00079-eng.htm (accessed 8 Dec 2020).

20 Sze S, Pan D, Gray LJ, et al. Ethnicity and clinical outcomes in COVID-19: A systematic review and meta-analysis. EClinicalMedicine 2020;29–30. doi:10.1016/j.eclinm.2020.100630

21 Koh WC, Naing L, Chaw L, et al. What do we know about SARS-CoV-2 transmission? A systematic review and meta-analysis of the secondary attack rate and associated risk factors. PLoS ONE 2020;15:e0240205. doi:10.1371/journal.pone.0240205

22 Madewell ZJ, Yang Y, Longini IM, et al. Household transmission of SARS-CoV-2: a systematic review and meta-analysis of secondary attack rate. MedRxiv 20164590 [Preprint]. August 1, 2020 [cited 2020 Nov 24] doi:10.1101/2020.07.29.20164590

23 Claveau J. The Canadian Housing Survey, 2018: core housing need of renter households living in social and affordable housing. 2020.https://www150.statcan.gc.ca/n1/en/pub/75f0002m/75f0002m2020003-eng.pdf?st=-9IHcxlK (accessed 24 Nov 2020).

24 Milan A, Laflamme N, Wong I. Diversity of grandparents living with their grandchildren. 2015.https://www150.statcan.gc.ca/n1/en/pub/75-006-x/2015001/article/14154-eng.pdf?st=NflKXopA (accessed 24 Nov 2020).

25 Chang S, Pierson E, Koh PW, et al. Mobility network models of COVID-19 explain inequities and inform reopening. Nature Published Online First: 10 November 2020. doi:10.1038/s41586-020-2923-3

26 McCormack G, Avery C, Spitzer AK-L, et al. Economic Vulnerability of Households With Essential Workers. JAMA 2020;324:388. doi:10.1001/jama.2020.11366

27 St-Denis X. Sociodemographic Determinants of Occupational Risks of Exposure to COVID-19 in Canada. Can Rev Sociol 2020;57:399–452. doi:10.1111/cars.12288

28 Messacar D, Morissette R, Deng Z. Inequality in the feasibility of working from home during and after COVID-19. 2020.https://www150.statcan.gc.ca/n1/pub/45-28-0001/2020001/article/00029-eng.htm (accessed 8 Dec 2020).

29 Hou F, Frank K, Schimmele C. Economic impact of COVID-19 among visible minority groups. https://www150.statcan.gc.ca/n1/pub/45-28-0001/2020001/article/00042-eng.htm (accessed 8 Dec 2020).

30 Turcotte M, Savage K. The contribution of immigrants and population groups designated as visible minorities to nurse aide, orderly and patient service associate occupations. https://www150.statcan.gc.ca/n1/pub/45-28-0001/2020001/article/00036-eng.htm (accessed 24 Nov 2020).

31 Guttmann A, Gandhi S, Wanigaratne S, et al. COVID-19 in Immigrants, Refugees and Other Newcomers in Ontario: Characteristics of Those Tested and Those Confirmed Positive, as of June 13, 2020. 2020.https://www.ices.on.ca/~/media/Files/Atlases-Reports/2020/COVID-19-in-Immigrants-Refugees-and-Other-Newcomers-in-Ontario/Full-Report.ashx (accessed 17 Nov 2020).

32 Berkowitz RL, Gao X, Michaels EK, et al. Structurally vulnerable neighbourhood environments and racial/ethnic COVID-19 inequities. Cities & Health 2020;:1–4. doi:10.1080/23748834.2020.1792069

33 Nestel S. Colour Coded Health Care: The impact of race and racism on Canadians’ Health. 2012.https://www.wellesleyinstitute.com/wp-content/uploads/2012/02/Colour-Coded-Health-Care-Sheryl-Nestel.pdf (accessed 9 Dec 2020).

34 Siddiqi AA, Wang S, Quinn K, et al. Racial Disparities in Access to Care Under Conditions of Universal Coverage. American Journal of Preventive Medicine 2016;50:220–5. doi:10.1016/j.amepre.2014.08.004

35 Veenstra G, Patterson AC. Black–White Health Inequalities in Canada. Journal of Immigrant and Minority Health 2016;18:51–7. doi:10.1007/s10903-014-0140-6

36 Chiu M, Maclagan LC, Tu JV, et al. Temporal trends in cardiovascular disease risk factors among white, South Asian, Chinese and black groups in Ontario, Canada, 2001 to 2012: a population-based study. BMJ Open 2015;5:e007232. doi:10.1136/bmjopen-2014-007232

37 Muñoz-Price LS, Nattinger AB, Rivera F, et al. Racial Disparities in Incidence and Outcomes Among Patients With COVID-19. JAMA Netw Open 2020;3:e2021892. doi:10.1001/jamanetworkopen.2020.21892

38 Kirby T. Evidence mounts on the disproportionate effect of COVID-19 on ethnic minorities. The Lancet Respiratory Medicine 2020;8:547–8. doi:10.1016/S2213-2600(20)30228-9

39 Sundaram ME, Calzavara A, Mishra S, et al. The Individual and Social Determinants of COVID-19 in Ontario, Canada: A Population-Wide Study. MedRxiv 20223792 [Preprint]. November 12, 2020 [cited 2021 Jan 08] doi:10.1101/2020.11.09.20223792

40 Buajitti E, Chiodo S, Rosella LC. Agreement between area- and individual-level income measures in a population-based cohort: Implications for population health research. SSM - Population Health 2020;10:100553. doi:10.1016/j.ssmph.2020.100553

